# Dose-response relationship between protein intake and muscle mass increase: a systematic review and meta-analysis of randomized controlled trials

**DOI:** 10.1101/2020.02.21.20026252

**Authors:** Ryoichi Tagawa, Daiki Watanabe, Kyoko Ito, Keisuke Ueda, Kyosuke Nakayama, Chiaki Sanbongi, Motohiko Miyachi

## Abstract

**Background:** Lean body mass (LBM) is essential for health; however, consensus regarding the effectiveness of protein interventions in increasing LBM is lacking.

**Objective:** Evaluate the dose-response relationship of the effects of protein on LBM.

**Data Sources:** PubMed and Ichushi-Web databases were searched. A manual search of the references of the literature included here and in other meta-analyses was conducted.

**Study Selection:** Randomized controlled trials evaluating the effect of protein intake on LBM were included.

**Data Extraction:** Two researchers independently screened the abstracts; five reviewed the full-texts.

**Results:** 5402 subjects from 105 articles were included. In the multivariate-spline model, the mean and corresponding 95% confidence intervals (CIs) for LBM increase for 0.1 g/kg body weight (BW)/day increment was 0.39 [95% CI, 0.36–0.41] kg and 0.12 [0.11–0.14] kg below and above total protein intake 1.3 g/kg BW/day, respectively.

**Conclusions:** Our findings suggest that slightly increasing current protein intake for several months by 0.1 g/kg BW/day may increase or maintain LBM in a dose-response manner from 0.5 to 3.5 g/kg BW/day.

## INTRODUCTION

Skeletal muscle, which is responsible for movement and activity, is the largest organ in the human body, accounting for 40% of the total body weight. Among young and middle-aged adults, decreased muscle mass increases the risk of chronic metabolic diseases such as type 2 diabetes and obesity.^1, 2^ Moreover, among the elderly, a progressive decrease in muscle mass (sarcopenia) with age is a risk factor for fractures, physical disabilities, and frailty.^3^ Accordingly, sustaining and increasing muscle mass is extremely important for the promotion and maintenance of health across all populations.^4^

Protein, an energy-producing nutrient, is a major component of skeletal muscle in living organisms, and is involved in the regulation of metabolism.^5, 6^ Decreased muscle mass may be accelerated by a decline in the assimilation response to insufficient protein intake.^7^ According to a meta-analysis of nitrogen delivery tests to evaluate the required amount of protein, protein requirement for adults was reported to be 0.66 g/kg body weight (BW)/day.^8^ However, although some randomized controlled trials (RCTs) reported increased skeletal muscle mass following intake of more than the required amount of protein,^9-15^ no consistent results have been demonstrated.^12^ The dose-response relationship between protein intake and muscle mass has been reported in a recent meta-analysis of RCTs.^14^ However, since this report mainly demonstrates the magnitude of effect in the dose-response curve, the dose-response relationship between increased muscle mass and protein intake cannot be estimated from confidence intervals (CI). Furthermore, only the studies that examined the effect of protein supplementation with RT were included for analysis, thus the effect of protein supplementation without RT was not considered. Consequently, the purpose of this systematic review and meta-analysis was to quantitatively evaluate the dose-response relationship between differences in various protein intake and an increase in lean body mass.

Protein and amino acid ingestion strongly stimulates muscle protein synthesis,^16^ and digested and absorbed proteins and amino acids also act as structural components of muscle hypertrophy.^5^ Additionally, resistance training (RT) also facilitates muscle protein synthesis and increases in muscle mass. We aimed 1) to evaluate the dose-response relationship in the effects of protein on lean body mass and 2) to assess these relationships considering the presence or absence of RT. The hypothesis in this study was that increased protein intake would result in an increase in muscle mass in a dose-response manner and that ingesting small amounts of protein, especially among a resistance-trained population, would be effective in increasing muscle mass. This study is the first meta-analysis examining the dose-response relationship between a wide range of protein intake levels and the increase in muscle mass, considering the presence or absence of RT. Findings of this study contain recommendations for appropriate amounts of protein intake required to sustain and improve muscle mass in a diverse population.

## METHODS

### Protocol

This study was registered in UMIN-CTR (Registration No. UMIN000039285).

### Reporting

This systematic review and meta-analysis was conducted in accordance with the PRISMA (Preferred Reporting Items for Systematic Reviews and Meta-Analyses) guidelines.^17^

### Data sources

A systematic review of published literature was conducted using the PubMed and Ichushi-Web (online database of academic articles in Japan) databases (last accessed on May 27, 2019). Results were limited to English and Japanese language RCTs. The combinations of search terms and search parameters are shown in Supplementary Table S1. Additionally, a manual search of the references of the literature included for this study and of that included in other meta-analyses was conducted.

### Study identification and data extraction

Two authors (R.T and K.N) independently screened titles and abstracts of all the search results, and eligibility was judged based on the eligibility criteria described later. Any disagreement regarding eligibility was resolved through deliberations. Articles judged to be potentially eligible in the primary screening, and articles for which no such decision could be made, were subjected to secondary screening to determine eligibility using the full-text version. Data on subjects’ attributes, intervention conditions, and the target outcome were extracted from the articles judged to be eligible during secondary screening. If a trial included more than one intervention group, each group was treated as a separate trial. Measurement results in the middle of the intervention period were excluded and only one result before and after the full intervention was utilized. When data required for creation of a forest plot could not be collected, the article’s corresponding author was contacted. In cases where numerical data were not available and responses on the inquiry could not be obtained from the corresponding author, but the data were available as graphs, numerical values were obtained using WebPlotDigitizer version 4.1 (Ankit Rohatgi, TX, USA).^18^ Five authors (K.I., K.U., R.T., C.S., and K.N.) conducted the secondary screening and data extraction, and two authors (K.I. and K.U.) conducted the verification.

### Eligibility

RCTs that studied the effects of protein intake on lean body mass (or fat-free mass if lean body mass was not available), where supplemental protein doses varied between study groups, were selected for analysis. The population, intervention, comparison, outcome, and setting (PICOS) criteria were used to define the research questions (Table 1). The target population was limited to subjects who did not have any serious illness (HIV, cancer, chronic renal failure, terminally ill, or disease that seriously affect physical activity). The protein intervention period was set as two weeks or more, which may be sufficient period for protein supplementation to enhance lean body mass increase^19^, so that we could collect all of the possible RCTs with various characteristics. Supplemental protein dose (g/day or g/kg BW/day) was set in advance of the intervention. Trials with inter-group differences in the amounts for interventions with muscle hypertrophy promoters (leucine, beta-hydroxy-beta-methylbutyrate, creatine, etc.) or vitamin D were excluded. When there was more than one control group, priority was given to the control group with equal energy intake and with larger differences in supplemental protein dose. Control groups with different conditions other than nutrition (such as exercise) were excluded.

**Table 1.** PICOS criteria for inclusion of studies.

| Parameter | Description |
| --- | --- |
| Population | Adult participants (not critically ill) |
| Intervention | Supplementary protein intake, supplemented for $\geq 2$ weeks |
| Comparator | Placebo or no intervention |
| Outcomes | Lean body mass or fat-free mass |
| Study design | Randomized controlled trial |

### Outcomes

When extracting data for muscle mass, muscle strength, and body fat mass as outcomes, the target of analysis in this systematic review and meta-analysis was lean body mass. For lean body mass, two values were recorded: lean body mass change in each group, and difference in lean body mass changes between an intervention group and a control group. The former was used to evaluate the effect of total protein intake calculated by supplemental protein dose and dietary protein intake in each group, and the latter was used to evaluate the effect of difference in supplemental protein doses between groups.

### Quality assessment

Two authors (K.I. and K.N.) independently evaluated the selected articles’ quality with Cochrane risk-of-bias tool.^20^ Disagreements on judgment were resolved through discussions with a third author (R.T.). To avoid bias, those articles with detailed descriptions that are usually excluded (compared to articles with less detailed descriptions), and all articles were evaluated, including the ones containing high-risk items.

### Statistical analysis

A meta-analysis was conducted on the effect of protein intake on lean body mass using mean change and standard deviation (SD) of change (SD_change_). In cases where SD_change_ was not reported, it was calculated using the equations given below.^20^ In cases where all data for SD before intervention (SD_baseline_), SD after intervention (SD_final_), and SD_change_ were available, correlation coefficient (Corr) was calculated using the following equation:

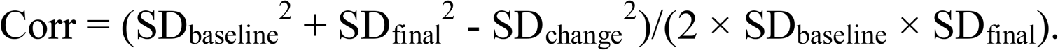

In cases where SD_change_ was unknown, but SD_baseline_ and SD_final_ were available, SD_change_ was calculated using the following equation:

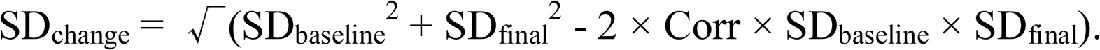

In cases where all the above data were not available, SD_change_ was obtained by contacting the corresponding author.

The effects of differences in supplementation doses on differences in lean body mass changes between groups were analyzed with point-estimation expressed by mean difference and 95% CI using a forest plot. To investigate the efect of RT, subgroup analyses were conducted. The analyses were performed using a random-effects model assuming that trial errors were included since the trials were selected using a wide range of conditions and were not limited by sex, age, and exercise conditions. Statistical heterogeneity was evaluated using the inconsistency index (*I*^2^) and *χ*^2^ test, but the entire analysis was performed even when heterogeneity was high, based on the assumption that corrections for confounding factors would be incorporated later. Publication bias was visually evaluated using a funnel plot analysis.

Moreover, unadjusted or multivariate-adjusted spline models were used to evaluate the dose-response relationship between protein intake (total protein intake in each group, or difference in supplemental protein doses between groups) and change in lean body mass (a value in each group or difference between groups). Multivariate analysis was verified in the following two models: in model 1, adjusted for age (continuous), sex (%male), intervention period (continuous), RT (yes or no); in model 2, in addition to the factors adjusted for in model 1, adjusted for weight change (continuous). The missing values of these covariates (of all the 105 selected trials; sex in eight cases, age in one case, weight change in 14 cases) were substituted with average values (for weight change only, missing values were substituted with 0) of all included trials. In Model 1, we adjusted for variables, which have been stratified variables used in a dietary reference intake (DRI)^21^, as confounders of the relationship between skeletal muscle and protein intake. The weight change was selected mediators of these relationships in model 2. Additionally, we performed the stratified-models analysis according to presence or absence of RT and the results were expressed as the effect size and 95% CI, with the former being calculated relative to the control group. The mean and the corresponding 95% CIs for lean body mass increase were estimated for a 0.1 g/kg BW/day increment in protein intake stratified by approximately 1.3 g/kgBW/day (< 1.3 or ≥ 1.3), which was the inflection point with the association between protein intake and FFM using a multivariate-spline model. In these analyses, when the 95% CI of the magnitude of effect did not straddle 0, it was estimated that p<0.05. When the 95% CI of the magnitude of effect straddled 0, it was estimated that p ≥0.05.

Statistical significance was considered when both sides were < 5%. Analyses were conducted using Review Manager (RevMan) version 5.3 (Nordic Cochrane Centre, Cochrane Collaboration, CPH, Denmark) and STATA MP, version 15.0 (StataCorp LP, College Station, TX, USA).

## RESULTS

### Study selection

The literature search results are shown in Figure 1. In the search conducted through May 27, 2019, 1700 potentially relevant articles were identified. Primary screening of titles and abstracts identified 295 articles that were either potentially eligible or for which no clear judgment could be made. Secondary screening using the full-text versions identified 149 eligible articles. The search was narrowed according to the target outcome for analysis in this meta-analysis; data for 105 articles, 138 intervention groups, and 5402 subjects were obtained. The data for intervention with RT were extracted from 53 articles, 72 intervention groups, and 2325 subjects. The data of intervention without RT were extracted from 56 articles, 66 intervention groups, and 3077 subjects. Four of the included articles reported on both interventions with and without RT. All 105 articles were used for a forest plot and spline models to evaluate the relationship between differences in supplemental protein doses and lean body mass change between groups, and 92 articles were used for spline models to evaluate the relationship between total protein intake and change in lean body mass in each group.

**Figure 1.**
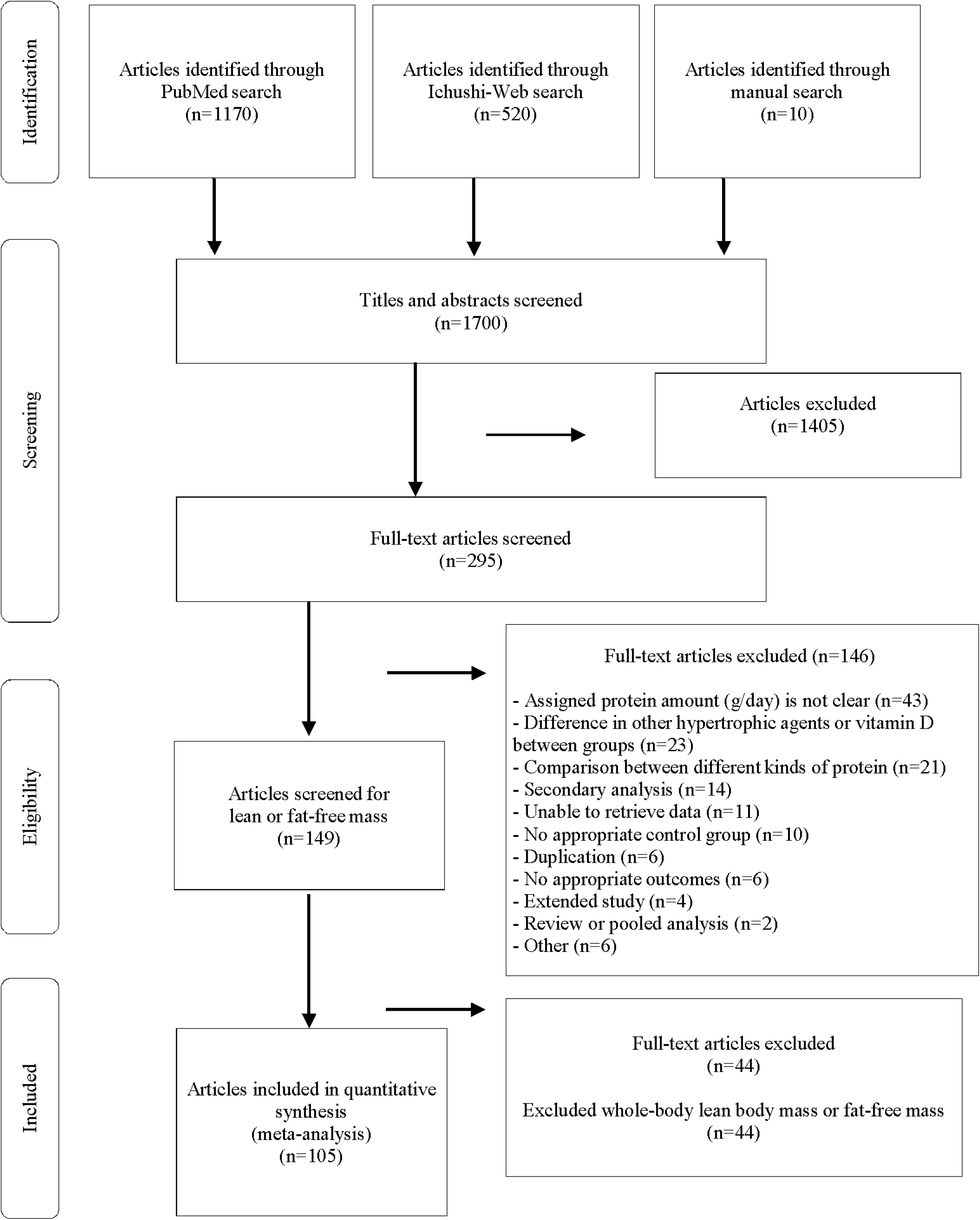
Flow diagram of the literature search process.

### Study characteristics

Supplementary Table S2-S5 summarizes the features of the 105 selected articles. Total protein intake ranged from 0.64 to 3.50 g/kg BW/day (mean ± SD: 1.58 ± 0.59 g/kg BW/day) in intervention groups and ranged from 0.52 to 2.00 g/kg BW/day (mean ± SD: 1.04 ± 0.35 g/kg BW/day) in control groups. There was a significant increase in total protein intake (g/day) in the intervention group (mean ± SD; range; p-value: 31 ± 27 g/day; −13 g/day to 135 g/day; p < 0.01) and a significant decrease in the control group (mean ± SD; range; p-value: −5 ± 15 g/day; −55 g/day to 47 g/day; p < 0.01) such that the change in total protein intake was significantly greater in the intervention group (p<0.01). Relative total protein intake (g/kg BW/day) significantly increased in the intervention group (pre: 1.13±0.33, post: 1.52±0.51, Δ: 0.38±0.33 g/kg BW/day, p<0.01) and significantly decreased in the control group (pre: 1.12±0.31, post: 1.06±0.33, Δ: −0.05±0.19 g/kg BW/day, p<0.01) such that there was a greater change in the intervention group (p<0.01). Differences in supplemental protein doses between an intervention group and a control group ranged from 0.06 to 2.38 g/kg BW/day (0.51 ± 0.37 g/kg BW/day). The RCTs comprised 68 trials evaluating lean body mass only, 35 trials evaluating fat-free mass only, and two trials evaluating both lean body mass and fat-free mass. There were 66 trials where protein supplementation was added to regular meals and 39 trials where the meal content itself was changed. The intervention period comprised a wide range, varying from 2 weeks to 18 months, with a mean of 19.8 weeks. Concerning energy balance, 41 trials entailed aggressive weight loss, two trials - aggressive weight gain, and 62 trials - neither. With regard to sex, 2459 subjects were female, 2422 male, and 530 subjects were unknown (data were not available). The mean age of the subjects ranged widely from 19-81 years in the trial groups, with an overall mean of 47.2 years.

### Risk of bias

Assessment of risk of bias is summarized in Supplementary Figure S1. High risk of bias included the following: blinding of participants and personnel in 58 trials, incomplete outcome data in seven trials, random sequence generation in three trials, and allocation concealment in three trials. Publication bias was not observed in the funnel plot (Supplementary Figure S2).

### Meta-analysis

Supplementary Figure S3 shows the forest plot consolidating the results of the trials for 138 intervention conditions stratified by presence or absence of RT. The results of subgroup analyses by supplemental doses with or without RT are summarized in Table 2. Protein supplementation was significantly effective in improving lean body mass in a wide range of doses with or without RT. As a whole, weighted average difference was 0.51 and 95% CI was 0.36 to 0.65 (p<0.01). For statistical heterogeneity, the *I*^2^ = 72%, and *χ*^2^ tests demonstrated statistical significance (p<0.01).

**Table 2.** Summary of the effect of protein supplementation on lean body mass change, stratified by the supplemental protein dose or stratified by the presence or absence of resistance training.

**change, stratified by the supplemental protein dose or stratified by the presence or absence of resistance training**
| Subgroup | Mean difference<br>(kg) | 95% CI<br>(kg) | Trials | Cumulative total<br>of subjects |
| --- | --- | --- | --- | --- |
| All trials | 0.51 | [0.36, 0.65] | 138 | 5866 |
| Less than 0.3 g/kg BW/day | 0.38 | [0.20, 0.55] | 40 | 1815 |
| 0.3 to 0.6 g/kg BW/day | 0.41 | [0.19, 0.63] | 56 | 2641 |
| 0.6 g/kg BW/day or more | 0.80 | [0.45, 1.14] | 42 | 1410 |
| With resistance training | 0.48 | [0.31, 0.65] | 72 | 2686 |
| Without resistance training | 0.53 | [0.36, 0.76] | 66 | 3180 |

*Abbreviation:* BW, body weight; CI, Confidence interval.

### Dose-response analyses with multivariate-adjusted spline models

The effect of total protein intake on lean body mass change in each group was analyzed with three spline models: unadjusted model, multivariate-adjusted model 1, or model 2 (Figure 2). In the analyses using unadjusted model or multivariate-adjusted model 1 (not including weight change), lean body mass change became incrementally greater with total protein intake in a whole range of intakes, with or without RT. When multivariate-adjusted model 1 was used, the mean and the corresponding 95% CIs for lean body mass increase for a 0.1 g/kg BW/day increment was 0.39 [95% CI, 0.36–0.41] and 0.12 [0.11–0.14] below total protein intake 1.3 g/kg BW/day and above total protein intake 1.3 g/kg BW/day, respectively. Positive correlations through a range of total protein intakes were also observed when stratifying the sample according to RT groups: [0.52–1.30 g/kg BW/day; mean, 0.06 (95% CI: 0.03 to 0.08) in RT and mean, 0.40 (95% CI: 0.37 to 0.43) in Non-RT, and 1.31–3.50/kg BW/day; 0.08 (95% CI: 0.06 to 0.09) in RT and 0.26 (95% CI: 0.23 to 0.29)]. In model 2 which added BW change as mediator to model 1, after exceeding total protein intake 1.3 g/kg BW/day, the effect on lean body mass change continued to rise with RT and declined without RT. The effect of differences in supplemental protein doses was analyzed in the same manner as above (Supplementary figure 4). Before and after differences in supplemental protein doses reached approximately 0.5 g/kg BW/day, the effect on lean body mass change declined and rose, respectively.

**Figure 2.**
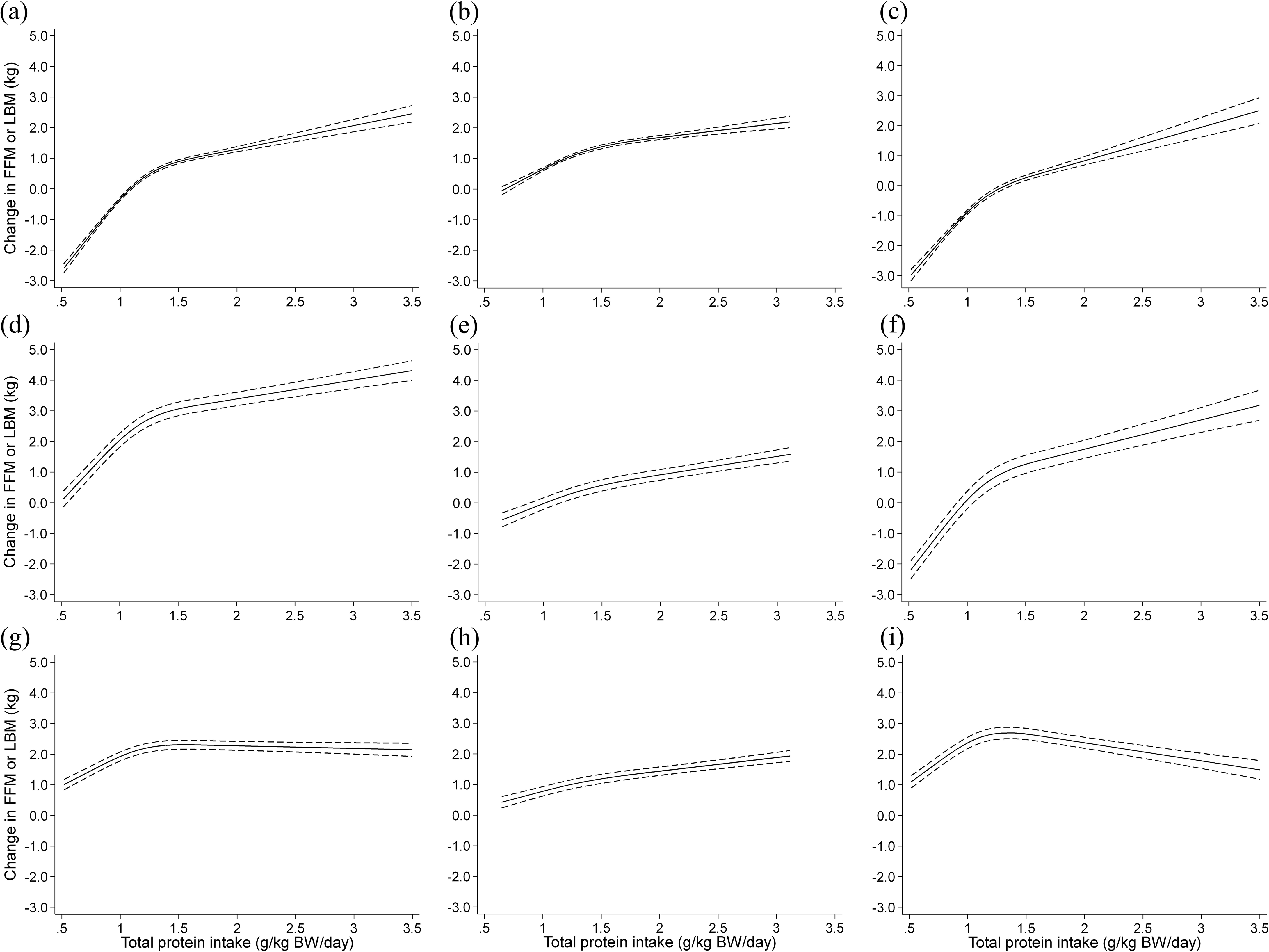
Dose-response relationship between total protein intake and change in lean body mass in each group. Spline curves illustrating the associations between total protein intake and change in lean body mass in each group in an unadjusted model (a, b and c for all trials, trials with resistance training and without resistance training, respectively), multivariate-adjusted model 1 (d, e and f for all trials, trials with resistance training and without resistance training, respectively) or model 2 (g, h and i for all trials, trials with resistance training and without resistance training, respectively). The solid line and dashed line represent the mean and 95% confidence intervals. Covariates of model 1 are age, sex, intervention period and resistance training. Covariates of model 2 are weight change in addition to the covariates of model 1.

## DISCUSSION

### Primary findings

The purpose of this study was to elucidate the dose-response relationship using a meta-analysis of RCTs investigating the effect of protein intake on lean body mass increase, considering the presence or absence of RT. The primary findings include: 1. the subgroup analyses of forest plot indicate that protein supplementation is significantly effective for increasing lean body mass with or without RT; 2. dose-response analyses with the multivariate-adjusted spline model indicate that total protein intake in a wide range (from 0.5 to 3.5 g/kg BW/day) is positively correlated with lean body mass increase. Slightly increasing the current protein intake by 0.1 g/kg BW/day may potentially increase or maintain the muscle mass: 3. the rate of increase in the effect of protein supplementation rapidly diminishes after exceeding 1.3 g/kg BW/day, and RT markedly suppresses this decline. These findings of this study contain recommendations for appropriate protein intake which is required to sustain and improve muscle mass in diverse populations and provide a better understanding of the influence of RT on the effect of protein intake.

### Effect of protein supplementation with or without RT

Several previous systematic reviews and meta-analyses of RCTs with RT report that protein supplementation has a significant positive effect on muscle mass^9, 10, 14^, but this meta-analysis has proved for the first time that protein supplementation is significantly effective without RT in a diverse population without specific conditions. Subgroup analysis according to RT demonstrated that no superior effects of protein supplementation were observed with RT (Table 2). It seems that RT has no synergistic effects, while it may have a simple additive effect^22, 23^.

### Effective dose

Subgroup analyses by supplemental protein doses demonstrate that protein supplementation of <0.3 g/kg BW/day (0.17 g/kg BW/day on average) is sufficient to significantly increase lean body mass. Furthermore, the multivariate-adjusted spline model reveals that total protein intake positively correlated with lean body mass change in a wide range (0.5 to 3.5 g/kg BW). Lean body mass increased by 0.39 (95% CI: 0.36–0.41) kg and 0.12 (95% CI: 0.11–0.14) kg per 0.1 g/kg BW/day increment in total protein intake below and above 1.3 g/kg BW/day, respectively. These results suggest that a small amount of protein supplementation promotes lean body mass increase. Daily addition of a high-protein food item such as an egg (6-8 g protein) or one cup (200 ml) of milk (6.8 g) per day to regular meals may increase muscle mass. These findings will also be informative for managing nutrition for people who have difficulty eating sufficient amounts, e.g. the elderly, people with dysphagia, patients, and people with financial constraints.

### Correlation between total protein intake and lean body mass change

Multivariate-adjusted spline model 1 (not including weight change) indicates that total protein intake and lean body mass change exhibited positive correlation in a wide range (from 0.5 to 3.5 g/kg BW/day), and the model 2 (including weight change) reveals that effect of total protein intake is especially efficient below 1.3 g/kg BW/day, indicating that increasing total protein intake within the range recommended by the dietary intake references^21^ leads to efficient and linear increases in lean body mass relative to body weight change. Considering the negative relationship with low doses between differences in supplementation doses and differences in lean body mass increases, total protein intake may be essential to accurately estimate dose-response of protein intake. Indeed, several previous reports^24-29^ indicate positive correlations between total protein intake and lean body mass in a diverse population.

### Effect of higher protein intake with or without RT

The rate of increase in the effect of protein supplementation rapidly diminishes when total protein intake exceeds 1.3 g/kg BW/day in the multivariate-adjusted spline model 2 (including weight change). This result suggests that the efficiency of conversion of ingested protein into lean body mass declines when protein is ingested in sufficient amounts or more. This finding is consistent with previous research using meta-regression between total protein intake and change in fat-free mass.^14^ Interestingly, this decline is markedly suppressed by RT, suggesting that RT may contribute to maintaining or improving the efficiency of protein anabolism. According to the national health and nutrition survey in Japan^30^, roughly 33% of Japanese adults may exceed 1.3 g/kg BW/day of total protein intake. Although estimated populations performing RT have been increasing steadily in Japan, currently only ~10% of Japanese adults perform RT once or more a week^31^. For health-oriented people who eat a lot of high-protein foods in daily meals, RT is strongly recommended to increase lean body mass.

### Strengths and limitations

This study has the following strengths. First, we included a large number of selected studies (trials) and subjects, approximately 2-3 times more than those in previous meta-analyses^9-15^. Second, dose-reactivity was described using a multivariate-adjusted spline model. Surprisingly, although numerous meta-analyses have been published to date, only a few analyzed dose-response relationships, despite major interests and concerns regarding the amount of protein intervention and its effect size. Third, this study is the first meta-analysis examining the dose-response relationship between a wide range of protein intake and lean body mass increase, considering the presence or absence of RT.

This study has some limitations. First, included studies were limited to those published in English and Japanese languages. Second, for articles in which the magnitude of effect on lean body mass was not mentioned in either the text or tables, the corresponding authors were contacted, but the response rate was low (seven out of 36 cases only). However, of the 29 articles where no response was obtained from the authors, 18 articles contained related graphs, and WebPlotDigitizer was used to extract data from these graphs. Thirdly, bias related to blinding was high. Double-blinded trials are difficult to perform in dietary protein interventions since it is necessary to provide meals with different content. However, as most studies of protein intervention are not double-blinded, excluding them may result in large deviations from the current status of protein intervention studies. Consequently, studies that were not double-blinded were also included in this meta-analysis.

### Perspectives

A future large-scale RCT is necessary to examine the dose-response relationship of multiple protein intervention amounts under the same conditions to more accurately elucidate the relationship between protein intervention amount and muscle mass increase. Only four of the 105 articles included in this meta-analysis examined the dose-response relationship under the same conditions. Moreover, intervention studies including subjects with issues due to severely insufficient protein intake (such as frailty and sarcopenia) are warranted. In developed and developing countries, sarcopenia and frailty among the elderly and kwashiorkor in young children, respectively, comprise some global health issues, which need to be resolved. Therefore, more well-designed and multi-faceted studies are necessary to further clarify the relationship between protein intake and muscle mass.

### Conclusion

This meta-analysis revealed that total protein intake enhances lean body mass increase in a dose-response manner in a wide range (0.5 to 3.5 g/kg BW). These results suggest that slightly increasing protein intake for several months, as by 0.1 g/kg BW/day may potentially increase or maintain muscle mass. The effect of protein intake on lean body mass increase relative to weight change rapidly diminishes after exceeding the intake of 1.3 g/kg BW/day, and RT markedly suppresses this decline. Therefore, both increasing protein intake and performing RT is recommended to efficiently augment lean body mass.

## Data Availability

All data referred to in the manuscript are completely saved and available.

## Acknowledgments

The authors would like to thank Aya Yoshimura, Yuri Saito, and Kae Yamazaki, researchers of Meiji Co., Ltd. for their support in data extraction.

## Author contributions

R.T. and D.W. are co-first authors and M.M. is the corresponding author. All authors contributed to the conception of research. M.M., R.T., and D.W. designed the research. R.T. and K.N. performed screening. K.I., K.U., R.T., C.S., and K.N. performed data selection and extraction. K.I. and K.N. performed quality assessment. M.M., D.W., and R.T. conducted the analyses and drafted the manuscript. All authors contributed to the critical review of the manuscript. M.M. had primary responsibility for the final content of the manuscript.

## Funding and Sponsorship

No external funding supported this work.

## Declaration of interest

R.T., K.I., K.U., K.N. and C.S. are employees of Meiji Co., Ltd.

## Supporting Information

The following Supporting Information is available through the online version of this article on the publisher’s website.

*Supplementary Table S1* **Search strategy for PubMed and Ichushi-Web**

*Supplementary Table S2* **Summary of characteristics of included studies**

*Supplementary Table S3* **Summary of nutrition surveys**

*Supplementary Table S4* **Summary of assigned protein amounts and differences between groups**

*Supplementary Table S5* **Summary of conditions of the studies’ interventions**

*Supplementary Figure S1* **Risk of bias assessment**

*Supplementary Figure S2* **Funnel plot of all the included studies for changes in lean body mass**

*Supplementary Figure S3* **Forest plot assessing the effect of protein supplementation on changes in lean body mass**

*Supplementary Figure S4* **Dose-response relationship between difference in supplemental protein doses and difference in lean body mass changes between groups**

## Notes

### Competing Interest Statement

The authors have declared no competing interest.

### Clinical Trial

UMIN000039285

### Funding Statement

No external funding was received.

